# Incidence and Severity of COVID-19 with the use of the MMR Vaccine before or after the COVID-19 Vaccine

**DOI:** 10.1101/2023.08.22.23294439

**Authors:** Edison Natal Fedrizzi, Alberto Trapani, Juliana Balbinot Reis Girondi, Aldanéa Norma de Souza Silvestrin, Maria Veronica Nunes, the MMRCoV Study Group

## Abstract

**Introduction:** The MMR vaccine has been shown by several studies over the years to have a potent effect on heterologous immunity. The reduction in mortality and respiratory and gastrointestinal diseases in childhood has been consolidated with recent studies demonstrating a better evolution of COVID-19 with the use of this vaccine. Stimulation of innate immunity by the MMR vaccine can be very useful, both used alone or in association with other vaccines, especially those for COVID-19.

**Objectives:** To evaluate the decrease in the incidence of infection or severity of COVID-19 with the use of the MMR vaccine before and after the use of specific vaccines against COVID-19.

**Methods:** This extension analysis followed 120 volunteer healthcare professionals aged 18 to 60 who received the MMR vaccine before the specific COVID-19 vaccine and 73 volunteers who used the MMR vaccine after the COVID-19 vaccine. Visits to the Research Center were carried out at an average interval of 4 weeks for 12 weeks. The diagnosis of COVID-19 was performed using the RT-PCR technique for the SARS-CoV-2 virus.

**Results:** The most used vaccine against COVID-19 was Coronavac in 59.1%. A total of 44 cases of COVID-19 were diagnosed (20% of the sample), the vast majority of which were mild cases (70.5%). There was no difference in the incidence and severity of COVID-19 in health professionals who used the MMR vaccine before or after the specific vaccine against the SARS-CoV-2 virus (Coronavirus or AstraZeneca).

**Conclusion:** The incidence and severity of COVID-19 does not differ with the use of the MMR vaccine before or after the specific vaccine against COVID-19.

## INTRODUCTION

When the measles vaccine was introduced in various countries of the world (1970s-80s), large reductions in infant mortality of 40% or more were observed (1-6). The protective effect was much greater than expected given the reduction in measles mortality. These observations suggested that the measles vaccine could have beneficial protective effects against infections other than measles (7). This non-specific immune response produces a potent protective barrier that can prevent cell invasion by another virus (1,8,9). A vaccine with a live and attenuated microorganism can stimulate the innate immune response, whose repeated exposure to the antigen (training of the innate immune response) (10-13) prolongs the time of action of this immune response (memory of the innate immune response) and, consequently, protection for other infections (heterologous immunity) for a longer time (14-17). However, this stimulus will provide protection for a limited period of time. Therefore, memory induction of innate immunity, although useful, is only an emergency action to protect against infections. Even if this effect is for a limited period of time, it can greatly contribute to reducing the spread of infection in the early stages of a pandemic and/or reducing the severity of the disease, preventing hospitalizations and deaths (18-20).

We recently published a clinical trial with more than 400 health professionals evaluating the action of the MMR vaccine on the evolution of COVID-19. The use of at least one dose of the MMR vaccine has shown a significant reduction in symptomatic COVID-19 by 42% and disease progression by 76% and 50% and 78% with two doses, respectively (21). This possible heterologous immunity of the MMR vaccine may be associated with the potent stimulation of innate immunity, leading to a decrease in the viral load by cellular immunity (2-6,18,22-25) and a possible action of humoral immunity by some similarities between the glycoproteins of the viruses of measles and rubella with SARS-CoV-2 (26,27) and the action of antibodies against mumps (28).

The aim of this study was to assess whether there is any difference in the incidence and evolution of COVID-19 with the use of the MMR vaccine before or after the specific vaccine against SARS-CoV-2, particularly Coronavac and AstraZeneca.

## METHODOLOGY

### Study Design and Participants

We recently published the effect of the MMR vaccine on the evolution of COVID-19 (MMRCoV Study) in 424 health professionals in the greater Florianópolis region (Santa Catarina – Brazil), evaluating heterologous immunity by stimulating innate memory immunity. During this 6-month analysis (through visit 7), starting the study in July 2020, we observed that the MMR vaccine reduced symptomatic COVID-19 and prevented its progression. It was further demonstrated that the MMR vaccine was completely safe in this cohort of men and women aged 18 to 60 years (21).

In February 2021, specific vaccination against COVID-19 began in Brazil and health professionals were a priority group at the beginning of the vaccination campaign. Therefore, we did not have the opportunity to assess the long-term immune response, since most volunteers received Coronavac, Pfizer or AstraZeneca vaccines during this period. As described in the Protocol, with confirmation of the effectiveness of the MMR vaccine against COVID-19, it was offered to all study volunteers who received placebo. One question that we seek to answer with the second stage of this study is whether there is any difference in the evolution of COVID-19 with the use of the MMR vaccine before or after the specific vaccines against the disease. As soon as the health professionals received the COVID-19 vaccine, the blinding of the study was opened and we revealed to which group they were allocated. The cohort that received placebo at the beginning of the study was vaccinated with the MMR vaccine, respecting the 4-week interval between the COVID-19 and MMR vaccines and 8 weeks for the two doses of the MMR vaccine.

In this second phase of the study, we followed 220 volunteers, 129 from the initial MMR cohort and 91 from the initial Placebo cohort. Of the 129 volunteers in the initial MMR cohort, nine did not receive any specific vaccine against COVID-19 and 93 were lost to follow-up, with 120 volunteers being evaluated for the effectiveness of the association of MMR and COVID-19 vaccines. Of the 91 volunteers in the initial Placebo cohort, one volunteer did not receive any vaccine (neither MMR nor COVID-19), 17 did not receive the MMR vaccine and 61 were lost to follow-up, with 73 volunteers being evaluated for the effectiveness of the combination of COVID-19 vaccines and MMR (Fig. 1).

**Fig. 1:**
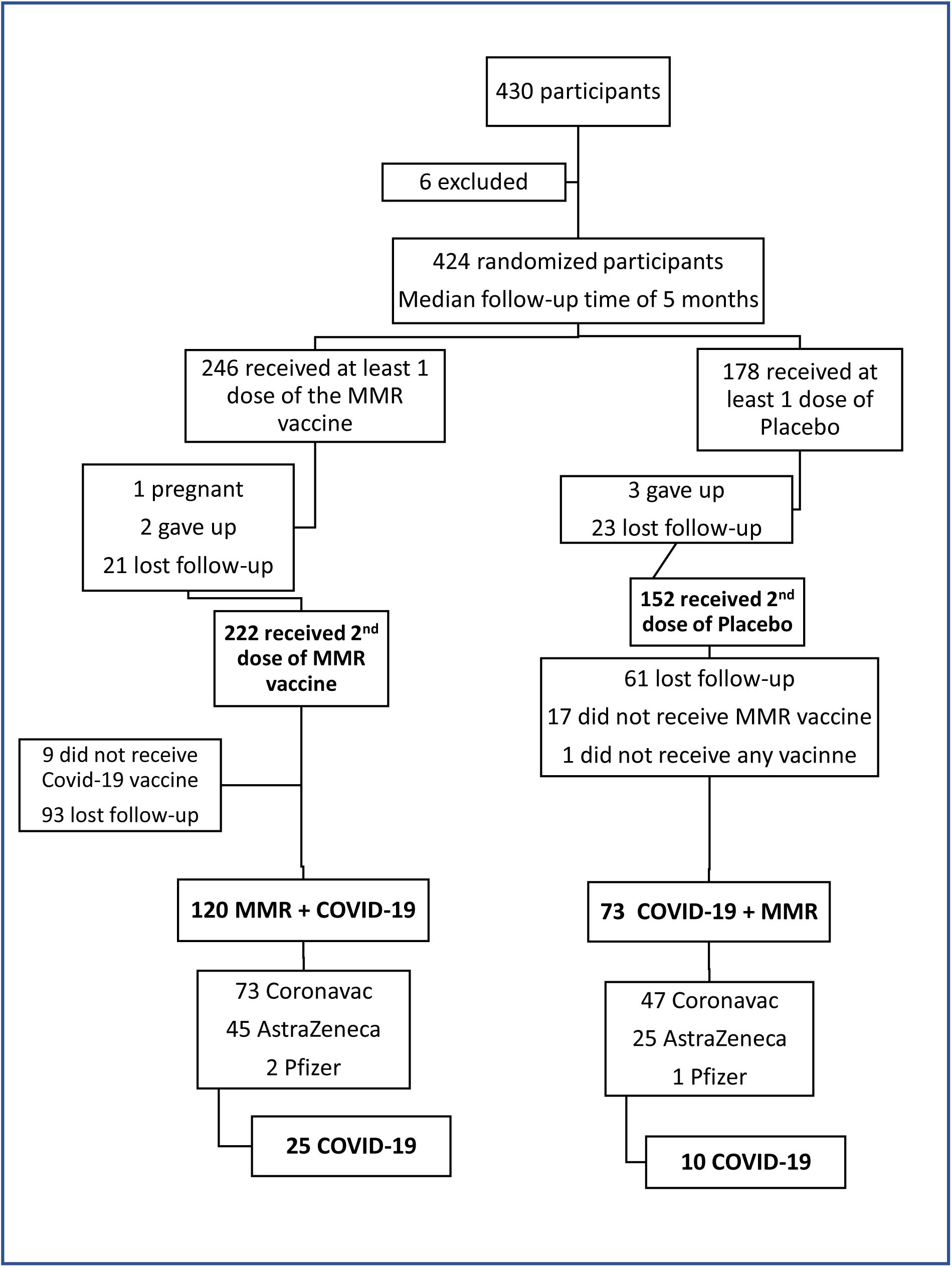
Study flowchart in Phase 1 and 2.

### Procedures

The second phase of the study involved monitoring 220 volunteers, especially those who used the MMR vaccine association (before or after) and specific vaccines against COVID-19. Thus, 193 volunteers (73 from the Placebo cohort and 120 from the MMR group) were followed for a period of 18 months (visit 17).

The volunteers visited the Research Center with an average interval of 4 weeks, where information was collected about their health, diagnosis of COVID-19, use of some medication and carried out the Rapid Test (IgG and IgM) for COVID-19 and collection oropharyngeal specimen for RT-PCR testing for SARS-CoV-2. The initial cohort that received placebo received two doses of the MMR vaccine, with an average interval of 8 weeks (Fig. 2).

**Fig. 2:**
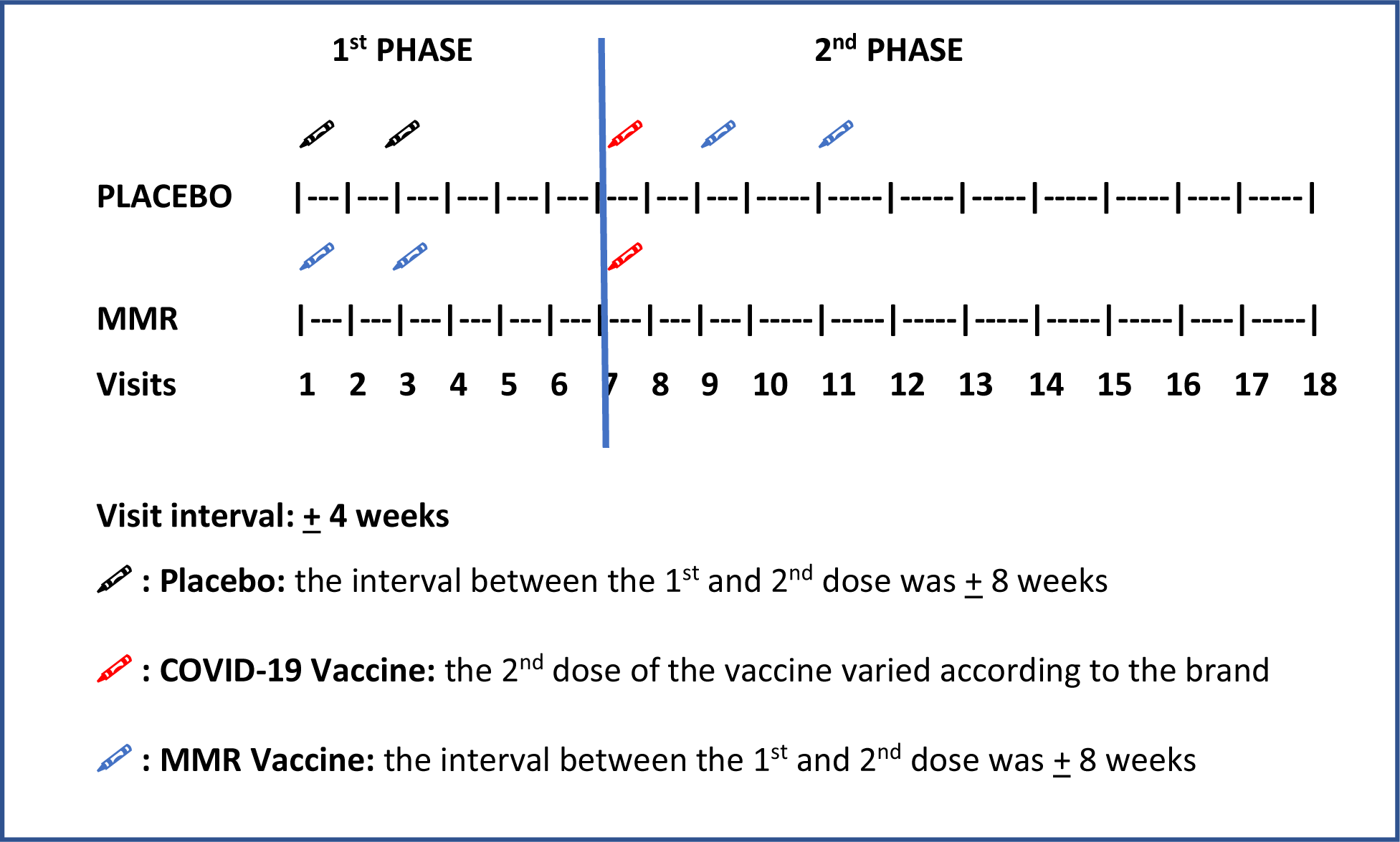
Procedures and visits in Phase 1 and 2 of the study.

A case, for the primary endpoint, was defined as a volunteer with a positive RT-PCR test for SARS-CoV-2, regardless of being symptomatic or not. To assess the severity of cases, we classified COVID-19 into 4 groups (29,30): 1) Asymptomatic: positive RT-PCR, absent symptoms; 2) Mild: positive RT-PCR, mild respiratory or clinical symptoms, with home follow-up; 3) Moderate: positive RT-PCR and respiratory or clinical symptoms that required non-symptomatic treatment such as anticoagulation, corticosteroid therapy and antibiotic therapy for pneumonia or hospitalization and 4) Severe: positive RT-PCR with respiratory symptoms or other complications that required admission to the Unit intensive care or evolved to death.

### Efficacy

The first outcome analyzed was the efficacy of the MMR + COVID-19 and COVID-19 + MMR vaccine combination in preventing RT-PCR-confirmed SARS-CoV-2 infection in participants with or without symptoms. The second outcome analyzed was the assessment of the severity of COVID-19 in these two cohorts.

### Statistical analysis

The efficacy of the MMR vaccine combination before or after the specific COVID-19 vaccine in preventing COVID-19 and/or reducing severity was demonstrated by calculating VE as (1-RR) * 100, where the relative risk (RR) was defined as the proportion of individuals with COVID-19 in Cohort 1 (MMR vaccine + COVID-19 vaccine) to the rate of individuals with COVID-19 in Cohort 2 (COVID-19 vaccine + MMR vaccine).

Efficacy would be demonstrated if the lower limit of the 95% confidence interval (CI) of vaccine combination efficacy was greater than 0%. The primary outcome analyzed was the incidence of COVID-19 and the secondary outcome, the severity of the disease in both cohorts.

## RESULTS

220 volunteers were followed, 129 from the initial MMR cohort and 91 from the Placebo cohort. Of the MMR cohort, 120 received a specific vaccine against COVID-19, while 9 volunteers did not want to receive the specific vaccine, having only the MMR at the beginning of the study.

In the Placebo cohort, 73 volunteers received the specific vaccine for COVID-19 and then the two doses of the MMR vaccine, respecting the interval between vaccines, according to the manufacturer. The interval between MMR vaccine doses was + 8 weeks. 17 volunteers did not receive the MMR vaccine and one did not receive either the COVID-19 vaccine or the MMR vaccine.

In this follow-up period, from visit 8 to 18 (with interval of + 4 weeks), 951 clinical appointments were performed, 396 in the Placebo cohort and 555 in the MMR cohort.

The COVID-19 vaccine most used by healthcare professionals in the study was Coronavac (130 volunteers – 59.1%), followed by AstraZeneca with 77 volunteers (35%).

44 cases of COVID-19 were diagnosed (20% of the sample), with the vast majority of mild cases (70.5%). See Table 1.

**Table 1:** Evaluation of volunteers and number of clinical appointments in the phase 2 of the MMRCoV Study in healthcare professionals.

|  | FREQUENCY |  |
| --- | --- | --- |
|  | n | % |
| VOLUNTEERS |  |  |
| MMR Cohort | 129 | 58.6 |
| - MMR + COVID-19 | 120 | 93.0 |
| - MMR | 09 | 7.0 |
| Placebo Cohort | 91 | 41.4 |
| - COVID-19 + MMR | 73 | 80.4 |
| - COVID-19 | 17 | 18.5 |
| - Placebo | 01 | 1.1 |
| TOTAL | 220 | 100 |
| CLINICAL APPOINTMENTS |  |  |
| Placebo Cohort | 396 | 41.6 |
| MMR Cohort | 555 | 58.4 |
| TOTAL | 951 | 100 |
| COVID-19 VACCINE |  |  |
| Coronavac | 130 | 59.1 |
| AstraZeneca | 77 | 35.0 |
| Pfizer | 03 | 1.4 |
| None | 10 | 4.5 |
| COVID-19 |  |  |
| Yes | 44 | 20.0 |
| No | 176 | 80.0 |
| COVID-19 SEVERITY |  |  |
| Asymptomatic | 12 | 27.3 |
| Mild | 31 | 70.5 |
| Moderate | 01 | 2.2 |

MMRCoV Study – HU/UFSC 220-2023.

Regarding the number of cases of COVID-19 in the 951 clinical appointments, we observed that there was no difference in the incidence of the disease in the two cohorts (MMR + COVID-19 and COVID-19 + MMR) (p=NS). See Table 2. The same was observed when we evaluated the 220 study participants separately, with 21% in the MMR + COVID-19 cohort and 16.4% in the COVID-19 + MMR cohort (p=NS). See Table 3.

**Table 2:** COVID-19 cases in the 951 clinical appointments of the phase 2 of the MMRCoV study in healthcare professionals.

| VACCINE | COVID-19 |  |  |  | TOTAL |  | p |
| --- | --- | --- | --- | --- | --- | --- | --- |
|  | YES |  | NO |  | n | % |  |
|  | n | % | n | % |  |  |  |
| MMR + COVID-19 | 27 | 5.1 | 504 | 94.9 | 531 | 55,8 | NS |
| COVID-19 + MMR | 10 | 4.7 | 202 | 95.3 | 212 | 22,3 |  |
| MMR | 03 | 13.0 | 20 | 87.0 | 23 | 2.4 |  |
| COVID-19 | 04 | 2.5 | 158 | 97.5 | 162 | 17.0 |  |
| PLACEBO | 00 | 0 | 23 | 100 | 23 | 2.5 |  |
| TOTAL | 44 | 5.5 | 907 | 94.5 | 951 | 100 |  |

MMRCov Study – HU/UFSC 220-2023.

**Table 3:** COVID-19 cases in the 220 volunteers in the phase 2 of the MMRCoV study in healthcare professionals.

| VACCINE | COVID-19 |  |  |  | TOTAL |  | p |
| --- | --- | --- | --- | --- | --- | --- | --- |
|  | YES |  | NO |  | n | % |  |
|  | n | % | n | % |  |  |  |
| MMR + COVID-19 | 25 | 21.0 | 95 | 79.0 | 120 | 54.5 | NS |
| COVID-19 + MMR | 12 | 16.4 | 61 | 83.6 | 73 | 33.2 |  |
| MMR | 03 | 33.3 | 06 | 66.7 | 09 | 4.1 |  |
| COVID-19 | 04 | 23.5 | 13 | 76.5 | 17 | 7.7 |  |
| PLACEBO | 0 | 0 | 01 | 100 | 01 | 0.5 |  |
| TOTAL | 44 | 20.0 | 176 | 80.0 | 220 | 100 |  |

MMRCov Study – HU/UFSC 220-2023.

When we evaluated the initial cohort that received the MMR vaccine and later the COVID-19 vaccine, we did not observe any statistical difference between the cases of COVID-19 and the type of vaccine used (Coronavirus 27.4% vs AstraZeneca 15.5%) (p =NS). The evaluation of the use of the Pfizer vaccine was not carried out by the very small number of users (two). The same occurred with the initially Placebo cohort that received the COVID-19 vaccine and then the MMR vaccine (Coronavirus 17.1% vs AstraZeneca 8%) (p=NS). See Table 4.

**Table 4:**
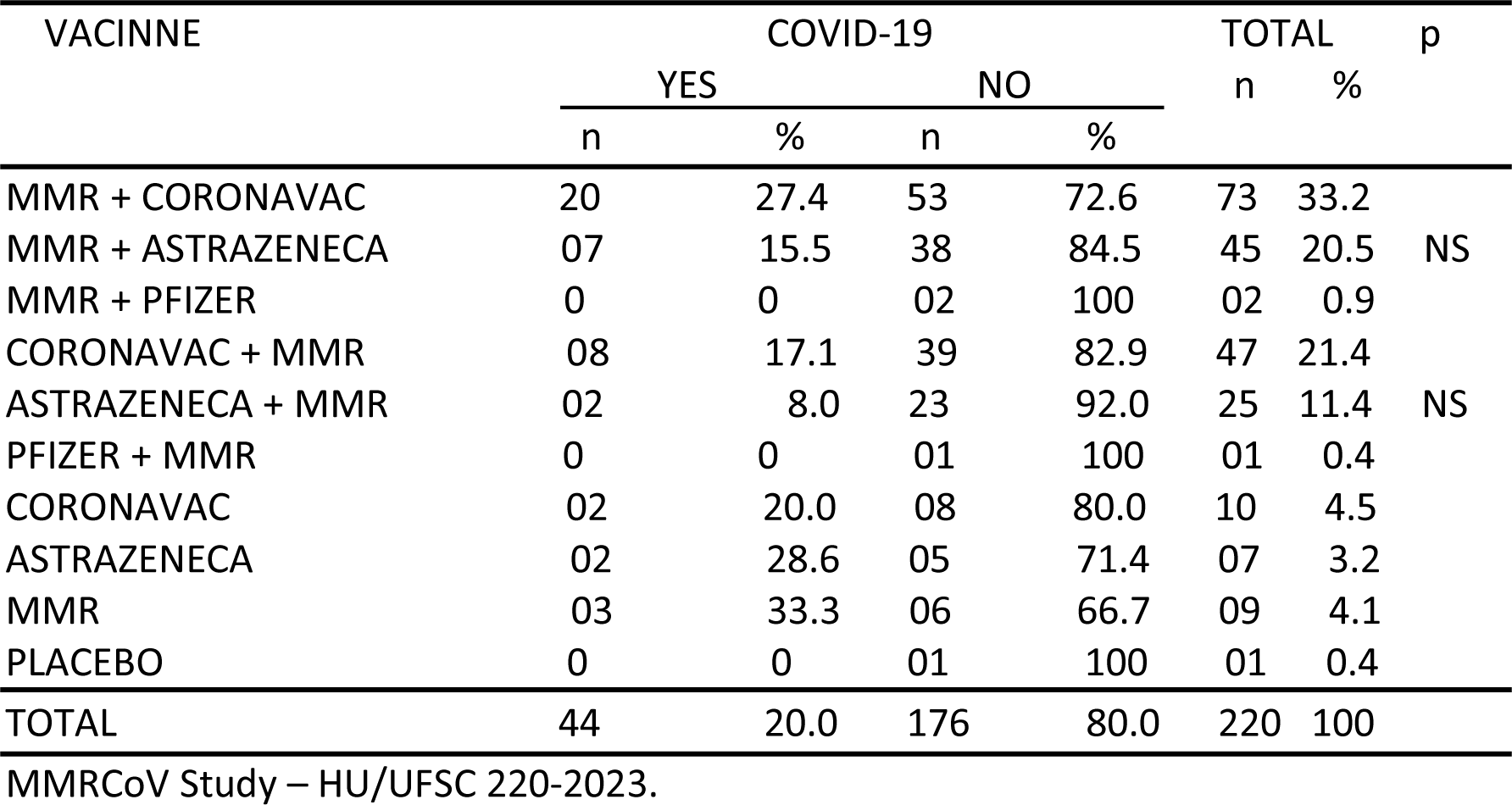
COVID-19 cases according to the type of COVID-19 vaccine and the time when the MMR vaccine was administered in the phase 2 of the MMRCoV study in healthcare professionals.

We had no cases of severe COVID-19 and only one case of moderate in the MMR + COVID-19 cohort. The vast majority of volunteers had mild cases (65.4%). Asymptomatic cases accounted for 32.7%. There was no statistically significant difference regarding the severity of COVID-19 and the use of the MMR vaccine before or after the specific COVID-19 vaccine (p=NS). See Table 5. When we evaluated the type of COVID-19 vaccine (Coronavac or AstraZeneca), we observed that there was no difference in the severity of COVID-19 between them and between the MMR + COVID-19 and COVIDE-19 + MMR cohorts (p= NS). See Table 6.

**Table 5:** Severity of COVID-19 according to the cohort that used the MMR Vaccine before and after the COVID-19 vaccine in the phase 2 of the MMRCoV study in healthcare professionals.

| VACCINE | COVID-19 |  |  |  |  |  | TOTAL |  | p |
| --- | --- | --- | --- | --- | --- | --- | --- | --- | --- |
|  | ASYMPTOMATIC |  | MILD |  | MODERATE |  | n | % |  |
|  | n | % | n | % | n | % |  |  |  |
| MMR + COVID-19 | 06 | 24.0 | 18 | 72.0 | 01 | 4.0 | 25 | 56.8 | NS |
| COVID-19 + MMR | 05 | 41.7 | 07 | 58.3 | 00 | 0 | 12 | 27.3 |  |
| MMR | 01 | 66.7 | 02 | 33.3 | 00 | 0 | 03 | 6.8 |  |
| COVID-19 | 00 | 44.4 | 04 | 55.6 | 00 | 0 | 04 | 9.1 |  |
| TOTAL | 12 | 32.7 | 31 | 65.4 | 01 | 1.9 | 44 | 100 |  |

MMRCov Study – HU/UFSC 220-2023.

**Table 6:**
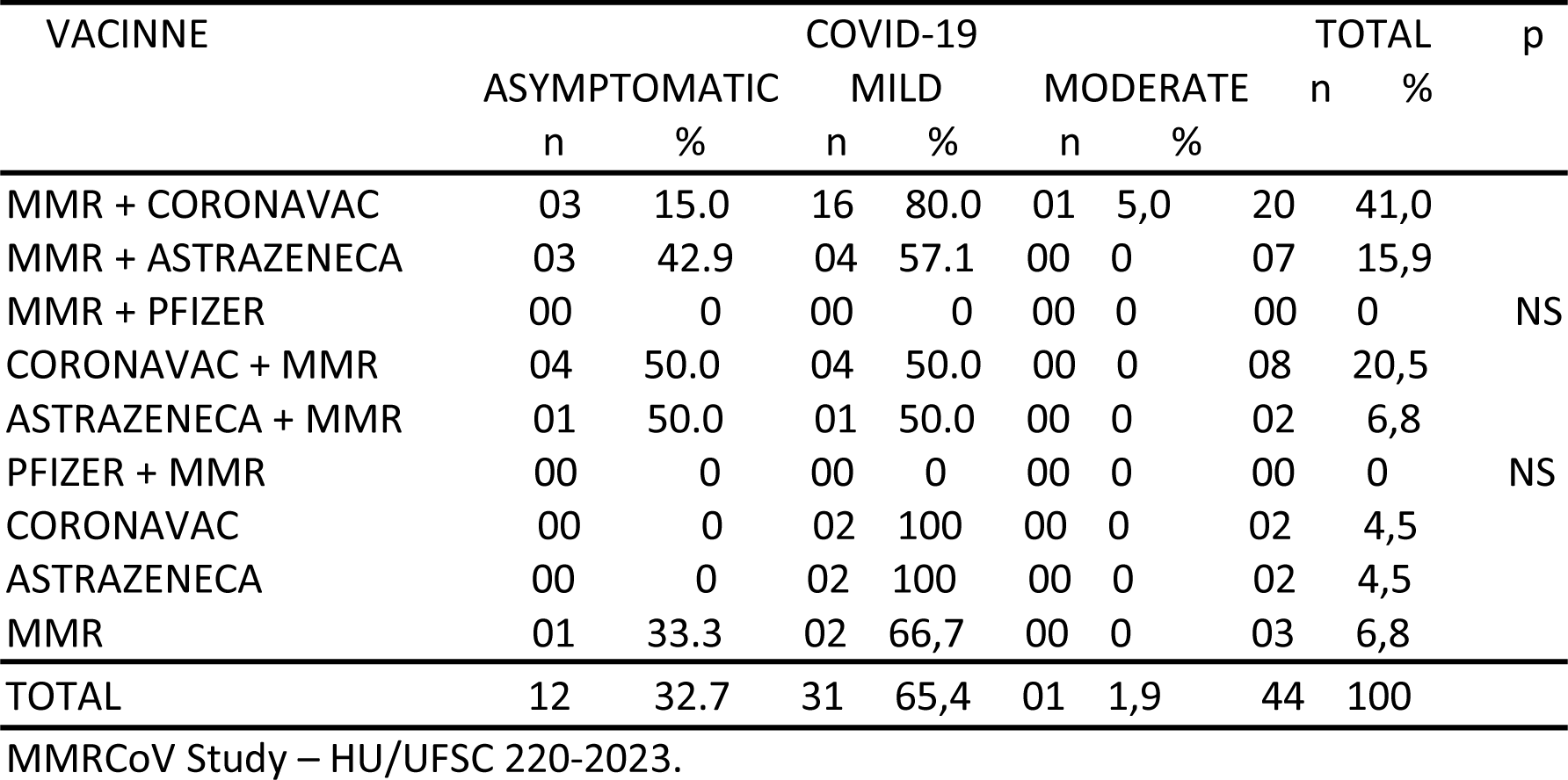
Severity of COVID-19 according to the COVID-19 vaccine and timing of the MMR vaccine in the phase 2 of the MMRCoV study in healthcare professionals.

## DISCUSSION

In a previous double-blind, randomized, placebo-controlled study (RCT), we demonstrated that the use of at least one dose of the MMR vaccine demonstrated a significant reduction of symptomatic COVID-19 by 42% and disease progression by 76%, confirming several previous studies that observed a reduction in infant mortality, respiratory and gastrointestinal infections, through heterologous immunity triggered by the stimulation of innate immunity by attenuated vaccines (1,7,12,14,17,18,31-41) and specifically with the MMR vaccine (2-6,18,22–25,30). Unfortunately, the duration of protection from heterologous immunity is still not well defined. In this study, the median follow-up was 5 months and it was not possible to assess the long-term effectiveness because the health professionals who volunteered for the study were immediately vaccinated with the specific vaccines against COVID-19. In this second phase of the study, we followed the volunteers for more 12 months, now evaluating whether the order of use of the MMR vaccine (before or after the COVID-19 vaccine) showed any difference in the incidence and evolution of the disease. This is the first clinical study to assess whether the order of use of the MMR vaccine (before or after the COVID-19 vaccine) makes any difference in disease progression.

Even after using specific vaccines against COVID-19, the percentage of positive cases was high, at 20%. This high percentage is probably justified because the participants are all from the health area and many work on the front lines. In a systematic review and meta-analysis of the prevalence of COVID-19 in health professionals diagnosed through RT-PCR, Gómez-Ochoa observed a high prevalence of infected people in the health area, of 11% (42).

Although the percentage of COVID-19 was lower (16%) in the cohort that received the MMR vaccine after the COVID-19 vaccine (compared to the 21% that received it before), this difference was not significant (p=NS). However, it is possible to infer that the heterologous immunity response is more robust in the cohort that received the MMR vaccine before the COVID-19 vaccine due to the greater interval between the two and there was no difference in the efficacy. Several studies have shown, however, that possibly the memory of innate immunity (stimulated by innate immunity training) can last for a year or more (17,20,43).

Most cases of COVID-19 have been mild or asymptomatic. Although asymptomatic COVID-19 was more frequent in the cohort that used the MMR vaccine after the COVID-19 vaccine (41.7% vs 24%), this difference was not statistically significant (p=NS). The same reasoning that we used in relation to the incidence of COVID-19 applies here. It can be postulated that when someone has a potent immune response and encounters the SARS-CoV-2 virus, a strong activation of local innate and adaptive immune responses keeps viremia in check, leading to low inflammation and systemic recovery. In contrast, in individuals with a defective immune response, due to old age or comorbidities, unrestricted viral replication leads to high concentrations of the virus, which in turn trigger hyperinflammation and serious complications such as severe acute respiratory syndrome and death (34,44). In an observational clinical study with 255 volunteers vaccinated with MMR vaccine, the authors observed that all 36 cases of COVID-19 in this group had only mild symptoms (20). Some justifications that demonstrate that the MMR vaccine could have some action against COVID-19 are: 1) potent stimulation of the innate immune response and its memory (10-13). Recent reports indicate that COVID-19 can suppress the innate immune response (45); 2) the MMR vaccine viruses and the coronavirus are RNA viruses and, perhaps, this could further benefit the stimulation of the innate immune response using this vaccine (14); 3) 30 amino acid sequence homology between the SARS-CoV-2 Spike glycoprotein (PDB:6VSB) and measles virus fusion glycoproteins (PDB:5YW_B) and rubella virus envelope glycoproteins (PDB:4ADG_A). These regions could be antigenic epitopes to stimulate humoral immunity, whose antibodies could protect against COVID-19. This could be one of the explanations for the lower involvement of children with COVID-19 (26,27); 4) the macrodomains of the SARS-CoV-2 virus and the rubella virus have an identical amino acid sequence in 29%. Hypothetically, these SARS-CoV-2 macrodomains could be recognized by rubella antibodies and provide a certain degree of protection. One study demonstrated that rubella antibody (IgG) levels increased in patients with COVID-19, similar to cases of a second rubella infection (27); 5) significant evidence of an inverse correlation between mumps antibody levels and COVID-19 severity, i.e., the higher the mumps IgG antibody concentration, the lower the COVID- 19 severity (28).

The incidence of COVID-19 was lower in health professionals who used the AstraZeneca COVID-19 vaccine, both after the MMR vaccine (15.5%) and before (8%), when compared to the Coronavac vaccine (27 .4% and 17.1%, respectively). The same was observed in relation to the severity of the disease, both for asymptomatic cases and for mild cases. However, these differences were not statistically significant (p=NS). Efficacy studies of vaccines against COVID-19 show very similar results, although prevention of symptomatic and severe disease has been shown to be greater for AstraZeneca (46).

This study had many limitations. The number of participants was small, with many losses to follow-up of the original study. The median follow-up was 12 months and this prevented us from having a more adequate assessment of the duration of action of the heterologous immunity. It was not possible to assess the cellular immune response and viral load of participants who had COVID-19. Certainly, if we had a control cohort of health professionals without the use of any vaccine, it would be possible to better assess long-term immunity. The specific vaccines used by the study participants were, in almost all cases, Coronavac and AstraZeneca and these results are not necessarily the same for other vaccines. The age range evaluated was 18-60 years and we cannot guarantee that the observed results may be the same in people above and below this age.

The results of this study demonstrated that the MMR vaccine before or after the COVID-19 vaccine had virtually the same results regarding the incidence and severity of COVID-19. As in the previous study it was proved that the MMR vaccine reduces the incidence of symptomatic COVID-19 and its evolution (21), it is quite possible that this action has been maintained throughout this study period and to say that the action of the heterologous immunity of the MMR vaccine is of at least 18 months. As this result is related to the stimulation of heterologous immunity, we can dazzle that the MMR vaccine could be useful in cases of new epidemics/pandemics as an emergency measure, until specific treatments or vaccines for each case are made available to the general population. Faced with the world scenario we are observing, especially with the new variants, it is very likely that the MMR vaccine, due to its mechanism of action, will continue to protect against new and future mutations of SARS-CoV-2. However, it is important to note that the MMR vaccine does not replace specific vaccines against COVID-19.

## CONCLUSION

There was no difference in the incidence and severity of COVID-19 in healthcare workers who used the MMR vaccine before or after the specific vaccine against the SARS-CoV-2 virus (Coronavirus or AstraZeneca) and possibly the heterologous immunity from the MMR vaccine lasted at least these 18 months of follow-up, being 6 months in the original study and 12 months in this extension study.

## Declaration of competing interests

All authors declare no competing interests.

## Data Availability

All data produced in the present work are contained in the manuscript

## Acknowledgments

We would like to thank the Federal University of Santa Catarina (UFSC) and Santa Catarina State Research and Innovation Foundation (FAPESC) for financing the study; University Hospital Prof Polydoro Ernani de São Thiago – HU/UFSC/EBSERH for the research site where the study was carried out; Fiocruz-Biomanguinhos Laboratory for the supply of vaccines; Santa Catarina State Health Secretariat (SES/SC) and Central Public Health Laboratory (LACEN) for conducting the RTPCR tests for SARS-CoV-2; Florianópolis Municipal Health Department (SMS/PMF), Health Center of São José Citty Hall (SMS/PMSJ) and UFSC Health Sciences Center (CCS/UFSC) for the supply of consumables for the study, and all volunteers who participated in this study.

## Author contributions

Edison Natal Fedrizzi: Conceptualization, Methodology, Writing original draft preparation, Project administration; Juliana Balbinot Reis Girondi: Supervision, Investigation; Aldanéa Norma de Souza Silvestrin: Supervision, Investigation; Alberto Trapani Junior: Investigation; Maria Veronica Nunes: Investigation, Funding acquisition;

## Role of the funding source

The funders of the study had no role in study design, data collection, data analysis, data interpretation, or writing of the report.

## Funding

This study was supported by the Federal University of Santa Catarina (UFSC), Fiocruz-Biomanguinhos Laboratory, Santa Catarina State Research and Innovation Foundation (FAPESC), Santa Catarina State Health Department (SES/SC), Health Center of Florianópolis Citty Hall (SMS/PMF), Health Center of São José Citty Hall (SMS/PMSJ) and UFSC Health Sciences Center (CCS/UFSC), Brazil.

## Notes

### Competing Interest Statement

The authors have declared no competing interest.

### Clinical Trial

Clinical Trials Registry: Brazilian Clinical Trials Registry (ReBEC RBR-2xd6dkj - https://ensaiosclinicos.gov.br/rg/RBR-2xd6dkj).

### Funding Statement

This study was supported by the Federal University of Santa Catarina, Fiocruz-Biomanguinhos Laboratory, Santa Catarina State Research and Innovation Foundation, Santa Catarina State Health Department, Health Center of Florianopolis Citty Hall, Health Center of Sao Jose Citty Hall and UFSC Health Sciences Center, Brazil.

### Author Declarations

Approval by the Ethics Committee: This study was approved by the Ethics Committee for Research with Human Beings of the UFSC (CEPSH UFSC) under registration CAAE: 52671521.7.0000.012.

